# Quantifying the waning of humoral immunity

**DOI:** 10.1101/2025.05.13.25327542

**Authors:** Ananya Saha, Hasan Ahmed, Cora Hirst, Katia Koelle, Andreas Handel, Peter Teunis, Rustom Antia

## Abstract

Immunological memory is a defining feature of immunity, yet surprisingly there is no consensus on how to quantitatively describe how antibody titers wane over time. A major problem is that the slow waning of antibody titers requires the collection of data for decades post-infection or vaccination. Our analysis of the largest existing dataset shows that a power-law model describes antibody waning better than other frequently used models. Our analysis suggests: (i) Protective levels of antibodies to many vaccine/virus antigens may be maintained for longer than previously estimated. (ii) The rate of waning of antibodies to protein toxoid vaccines such as tetanus may be similar to those elicited by live virus infections. (iii) The long-term waning of antibodies can be estimated from data for a much shorter time-frame of about 1-3 years following immunization, suggesting that using a power-law analysis could allow rapid estimation for the waning of immunity to new vaccines.

---

The phenomenon of protection following infection is a central feature of the adaptive immune response and was documented by Thucydides who noted that those who recovered during a plague in Athens around 430BC could take care of the sick [1]. The potential longevity of this protection was noted by the Danish physician Panum who noted that those who contracted measles in an outbreak in the Faroe islands in 1781 did not get ill during the next measles epidemic 65 years later [2]. However despite substantial advances in our understanding of adaptive immune responses we still do not have a quantitative understanding of how rapidly immunity wanes [3, 4, 5]. This is not only a fascinating question in basic immunology, but also, an accurate description of waning has potential utility for design and implementation of vaccination regimes [6, 7]. Here, we focus on the waning of systemic IgG antibody responses.

Currently there is no consensus about how to quantitatively describe the pattern of waning and a number of different models have been used. Since antibody molecules have a short half life of weeks, the waning of antibody titers over a timescale of years reflects a loss in the number of antibody secreting plasma cells. Based on the simple assumption that plasma cells have a constant probability of dying per unit time the waning of antibody titers has been modeled as an exponential decay. However the exponential rate of waning is relatively rapid for the first few years and slower in subsequent decades [8, 9, 10, 11]. Consequently, the simple exponential decay has been extended to a two phase exponential decay model [8, 12, 13, 14, 15, 16, 17, 18, 19]. Another assumption is to consider that plasma cells generated following infection have a continuous distribution of probabilities for death, and this gives rise to waning that is described by a power-law [20, 21, 22, 23, 24]. The power-law decay also captures the rapid initial waning of antibody titers followed by slower waning.

There are a number of reasons for the lack of consensus between studies. A major problem is the limited amount of data available. Most studies measure the waning of antibodies do so for a relatively short duration, typically less than a decade, compared with the timescales for loss of protection which can be many decades [18, 25, 26, 27, 28]. Moreover studies typically focus on a single vaccine or infection, and furthermore different studies calculate antibody titers in different ways (e.g. AUC vs midpoint-titers), making it hard to compare results of different studies. To our knowledge only one study, that of Amanna et al, has data on the waning of antibodies to multiple virus and vaccine antigens over an extended period of more than a decade [8]. Earlier analysis of this data used an exponential decay model for the waning of antibodies at times greater than 3 years after infection or vaccination (i.e. after the initial rapid waning had slowed) [8, 13]. In this paper we revisit this dataset critically including and focusing on data in the first years after vaccination that had been ignored in the earlier analysis. This allows us to address the following questions: (i) What functional form best describes the waning of antibodies (e.g. is it linear, exponential or a power-function)? (ii) How does the waning of antibody titers depend on the type of infection or vaccination? (iii) Over what timeframe do we need to collect data to accurately estimate the waning of antibodies over the long-term?

## 1 Star Methods

We analyze longitudinal data from [8] which follows the titer of antibody responses to a number of vaccine and viral antigens in a group of 45 individuals. Serum samples from these individuals were collected as part of a serological study of individuals working in close proximity to nonhuman primates, and antibodies to a panel of virus and vaccine antigens were measured using ELISA assays as described in detail in [8]. ELISA titers do not give absolute antibody concentrations and we define the normalized antibody titer as the titer relative to the threshold of protection which was taken from the literature.

Fitting to a power-law model requires knowing the time of infection. We identify infections in the time series as a spike in antibody titer which was over 2 fold. The full dataset is shown in the Supplementary Text (Fig S1 in Supplementary Text). We modeled the waning of antibodies following infection or immunization at time *t* = 0 which we approximated as the midpoint between the time-points immediately prior to and following the serospike. We considered three models: a simple exponential model, a bi-phasic exponential model and a power-law model.

In the exponential waning model, the antibody titer is modeled as

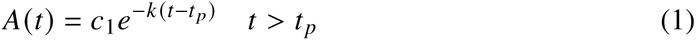

where *k* is the rate of waning per year and *c*_1_ is the magnitude of the response at time *t_p_* which is at or after the peak of the response. The half-life for waning of antibodies is given by *T*_1/2_ = ln(2)/*k*. The biphasic exponential model has two phases, a rapid initial exponential decay prior to *t* = 3 years and subsequent slower exponential decay at times greater than 3 years after infection. The *t* = 3 year cutoff was chosen for consistency with the timeframe for analysis of antibody waning in earlier studies [8, 13].

For the power-law decay, antibody titer is modeled as

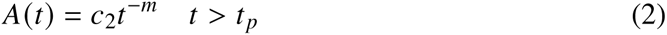

where *m* is the power-law constant and *c*_2_ is the antibody titer at *t* = 1 year after immunization or infection, and the peak titer 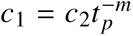. Since the data is not sufficiently detailed to accurately measure *t_p_* we take *c*_2_ as a rough measure of magnitude of the response. The power-law constant *m* equals the rate of waning per log time, which equals the slope of a plot of the log of the antibody titer versus the log of time. In the case of power-law waning, the rate of waning *k* (*t*) decreases with time as *k* (*t*) = *m*/*t*, and the time it takes for antibody levels to fall to half their current value is given by *T*_1/2_ = *t* (2^1/^*^m^* − 1).

To estimate the parameters of the exponential models and the power-law model, we used a linear mixed effects modeling framework, relying on the *lmer*() function in R [29] as described previously for exponential waning models [8, 13]. For the power-law model, we fitted log_10_(*A*)*_i j_* over log_10_(*t*)*_i j_* as,

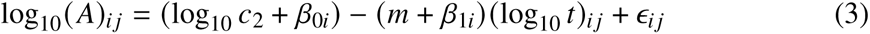

where the indices *i* and *j* are for the individual and time-points, *β*_0,_*_i_* and *β*_1,*i*_ are the random effects for the magnitude and slope, respectively, and *ɛ* is the error. Statistical support for comparing the fits of the different models to data was determined using the Akaike Information Criterion (AIC) [30]. Specifically, we determined the difference in AIC (Δ*AIC*) for the models, which indicates the measure of support for the model with the lower AIC. Larger values of Δ*AIC* indicate increasing support with 4 < Δ*AIC* < 10 indicating some support and Δ*AIC* > 10 indicating strong support.

## 2 Results

### Waning of antibody titers follows a power-law model

We begin by examining the waning of antibodies following vaccination against tetanus for which we have the most data. In Fig 1 and Fig S2 (in Supplementary Text) we plot the titer of tetanus-specific antibodies on a log scale versus time on a linear scale, so the fits to a simple exponential models is a straight line. Visually we see that the simple exponential model (dotted lines) does not fit the entire time-series well as it does not capture the decline in the rate of antibody waning over time. Both the bi-phasic exponential model (dashed lines) and the power-law model (solid lines) do much better, with the power-law visually appearing to be slightly better than the biphasic exponential waning model. This is confirmed by statistical analysis using AIC values. As shown in Table 1 the power-law model fits the data much better (ΔAIC=64) than the bi-phasic exponential waning model.

**Figure 1:**
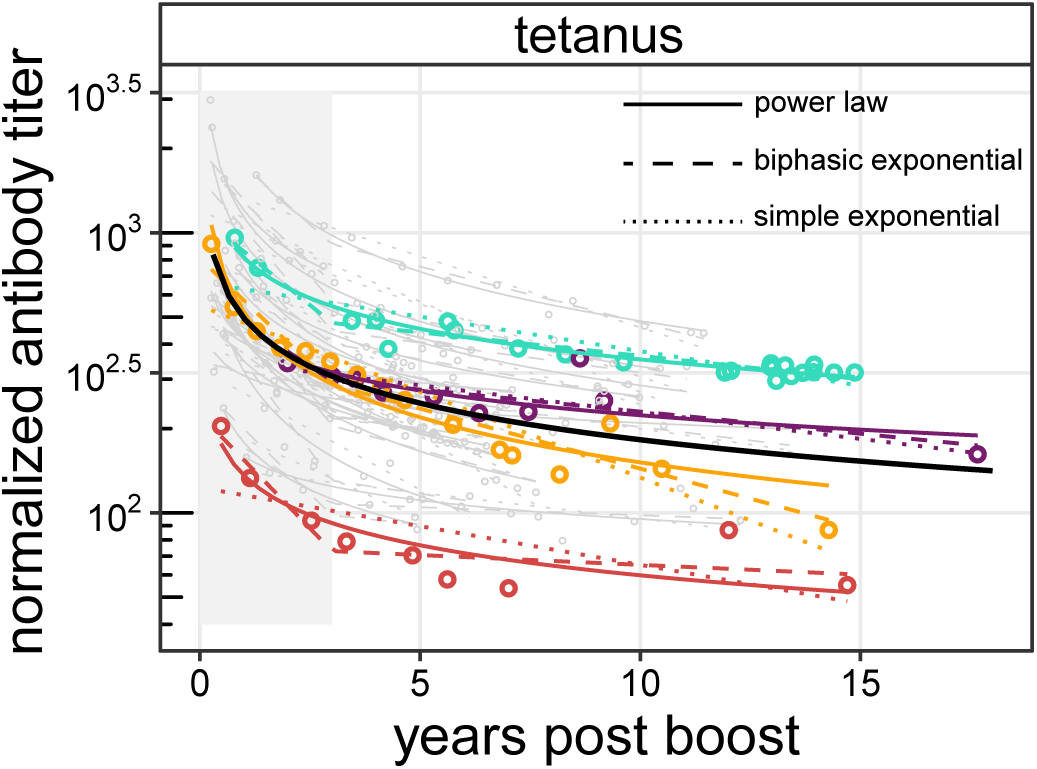
Waning of antibodies to tetanus follows a power-law. We plot the normalized antibody titer (i.e. scaled with respect to the threshold for protection) to tetanus as a function of time after vaccination. Plots for four individuals with the longest timeseries in color. Solid lines are fits to the power-law model, dotted and dashed lines are fits to the simple and bi-phasic exponential models respectively. The gray area demarcates the “early” from “late” phase for the bi-phasic waning model used by earlier studies [8, 13]. Statistical analysis indicates that the power-law model fits substantially better than either exponential model ΔAIC > 50 (see Table 1). The solid black line represents the estimated average for the antibody waning for the power-law model.

**Table 1:** AIC values for the fits of the simple exponential, bi-phasic exponential and power-law models for the waning of antibodies to different vaccine and virus antigens. The Δ*AIC* values, shown in red, indicate the degree of support for the best model compared with the next best. Δ*AIC* < 4 indicating NS, 4 < Δ*AIC* < 10 indicates some support; and Δ*AIC* > 10 indicating strong support. Finally, we estimated the waning of antibodies simultaneously to all of the vaccines and viruses listed in Table 1 and found that the power-law model fit these data substantially better than either the simple exponential model or the biphasic exponential model.

| Antigen | Model |  |  |
| --- | --- | --- | --- |
|  | simple exp | bi-phasic | power-law |
| Tetanus | -369.5 | -515.8 | -579.9 (64.1) |
| Diphtheria | -181.7 | -243.2 | -281.8 (38.6) |
| Vaccinia | 25.2 | NA | -89.1 (114.1) |
| VZV | -80.6 | -133.2 (1.2) | -132 |
| Measles | -16 | -24.9 | -27.3 (2.4) |
| Overall | -335.1 | -854.5 | -1008.5 (154) |

The waning of antibody titers to other virus and vaccine antigens also follows a power-law. In Fig 2 we show the fits for the waning of antibody titers to the power-law model for all vaccinations and infections (the fits to the exponential and bi-phasic exponential models are shown in Fig S3). In Table 1 we compare the performance of the power-law and exponential models by comparing the AIC of the fits of the different models to the data shown in Fig 2. Overall, we see that the power-law gives a much better fit to the data with Δ*AIC* > 150. For tetanus and diphtheria, for which we have the most data, the Δ*AIC* values of 64 and 38, respectively, indicate strong support for the power-law model. For measles and VZV, for which we have much less data and somewhat shorter timeseries, the power-law and bi-phasic model have comparable AIC values (Δ*AIC* < 4). For vaccinia, the power-law model is much better than the simple exponential and there is not sufficient data at times greater than three years to allow fitting to the bi-phasic model.

**Figure 2:**
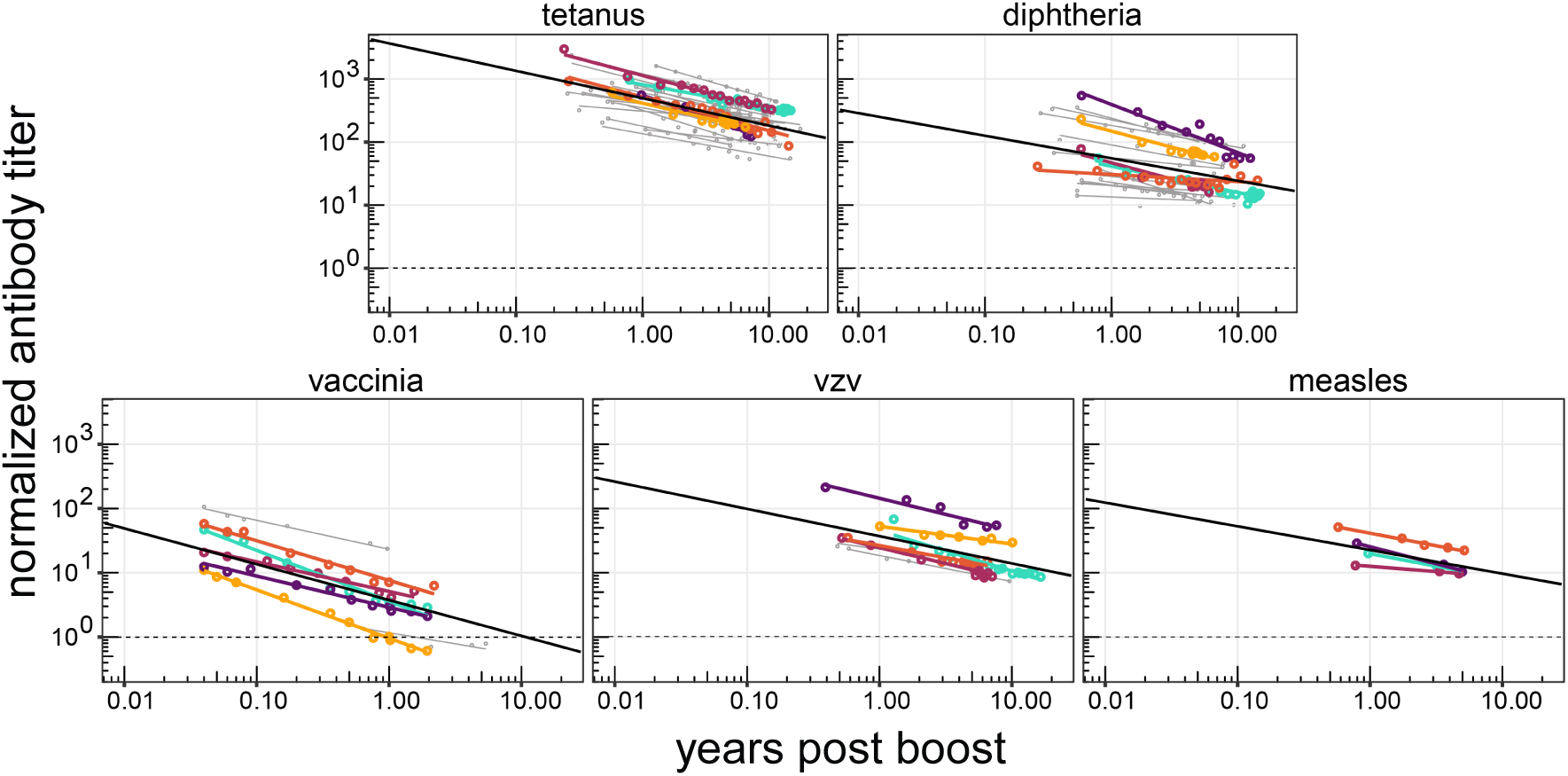
Waning of antibodies to all infections follows a power-law. The log of normalized antibody titers (scaled with respect to the threshold for protection) declines linearly with the log of time after vaccination or infection. As in Fig 1 we highlight four representative individuals for each infection in color and the black line in each panel represents the average decay. The dashed horizontal line corresponds to the loss of protection.

### Quantifying antibody waning to different viruses and vaccines

The power-law model has two parameters that describe the waning of antibody titers: *c*_2_ the antibody titer at 1 year post infection and *m* the power-law coefficient. In Fig 3 we compare the estimates of these two parameters for different virus and vaccine antigens. Fig 3A shows that different infections and immunizations result in very different titers (*c*_2_) shortly after infection or vaccination. This is consistent with results from earlier analysis of this dataset [8, 13].

**Figure 3:**
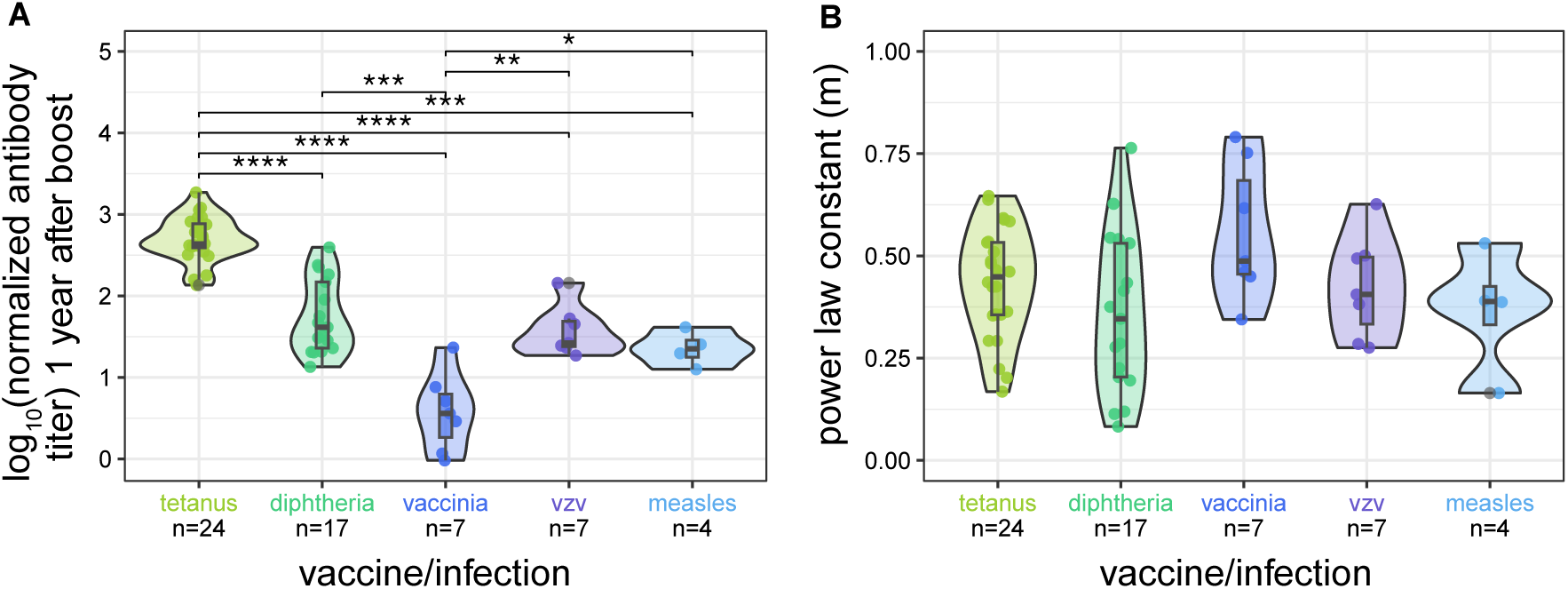
Parameters for the waning of immunity to different viruses and vaccines. In panel **(A)** we plot the estimates for the relative magnitude of the response (*c*_2_ in eqn 3) compared with the protective titer at *t* = 1. We find considerable variation in the magnitude of the responses to the different antigens at this time. In panel **(B)** we plot our estimates for the decay parameter *m* that describes the waning of antibodies to the different viruses and vaccines antigens. We find *m* is not significantly different for protein vaccines and virus infections.

Intriguingly, our results also indicate that the power-law coefficient (*m*) estimates for the waning of immunity are broadly similar across the different vaccines and virus antigens that we analyzed (Fig 3B). This result differs from the conclusions of previous analyses, which suggested, based on fits of exponential decay to data for *t* > 3 years after immunization, that antibody titers to tetanus and diphtheria wane much more rapidly (half-life estimates between 10-20 years) compared to antibodies following infection or immunization with live viruses such as vaccinia and measles (half-live estimates between 100-1000 years respectively) [8]. We now consider the cause of the differences between results of the earlier and current analysis.

The power-law model suggests one of the factors responsible for the much more rapid waning to protein / toxoid vaccines compared with live virus infections is the time at which waning was measured. A power-law predicts that the exponential rate of waning *k* decreases over time as *k* (*t*) = *m*/*t*. The earlier estimates for the rate of waning *k* for toxoid vaccines (tetanus and diphtheria) were relatively high because they were estimated from data relatively soon (in the first 15 years) post vaccination, while that for live virus infections (measles, vaccinia and VZV) were estimated in adults, about 40-50 years following childhood infection or vaccination. Consequently the rate of antibody waning would be expected to fall about 5.5 fold between year 8 (when it was typically estimated for tetanus and diphtheria) and year 45 (when it was typically estimated for the live virus infections). Applying this, antibodies to tetanus and diphtheria would have half-lives of 72 and 88 years, respectively if they were measured at 45 years post-vaccination. This is much closer to the estimates for the half-lives for the waning of antibodies to VZV and vaccinia (50 and 92 years, respectively) estimated in the earlier studies. In Table S1 (in Supplementary Text) we show that when we take into account the time at which waning is estimated there is considerably less difference between the earlier estimates using an exponential models and our estimates using the power-law model.

### A power-law model predicts longer times to loss of protective immunity

We now consider how our analysis changes estimates for the time to loss of protective immunity. We define the time to loss of protective immunity as the time for antibody titers to wane to the level defined to be protective for that vaccine or virus antigen. The levels of antibodies that are protective are taken from the literature [31, 32, 33, 34, 35]. In Fig 4A, we show projections for antibody titer using the parameterized power-law model and the parameterized bi-phasic exponential model for tetanus. In Fig 4B, we plot the proportion of a cohort that would be immune to tetanus infection as a function of time since vaccination. We find that virtually all individuals in such a cohort would have protective levels of antibodies to tetanus shortly following a boost. However, the rate of waning varies considerably depending on the underlying model. In particular, the power-law model predicts that all individuals in this cohort would have antibody titers exceeding the threshold for protection for tetanus for over 100 years (which exceeds a typical human lifespan). In contrast, the bi-phasic exponential model, and even more so, the single exponential model, both predict that the proportion of the cohort that is immune to tetanus decreases more rapidly. Our finding that the time to loss of immunity is substantially longer under a power-law model than under an exponential models holds true also for other viruses and vaccines. In Fig 4C, we plot the proportion of a cohort that would be immune to diphtheria, vaccinia, and VZV infection as a function of time since vaccination. Consistent with our results for tetanus, the proportion of individuals who are expected to be immune is substantially higher under the power-law model than either exponential model.

**Figure 4:**
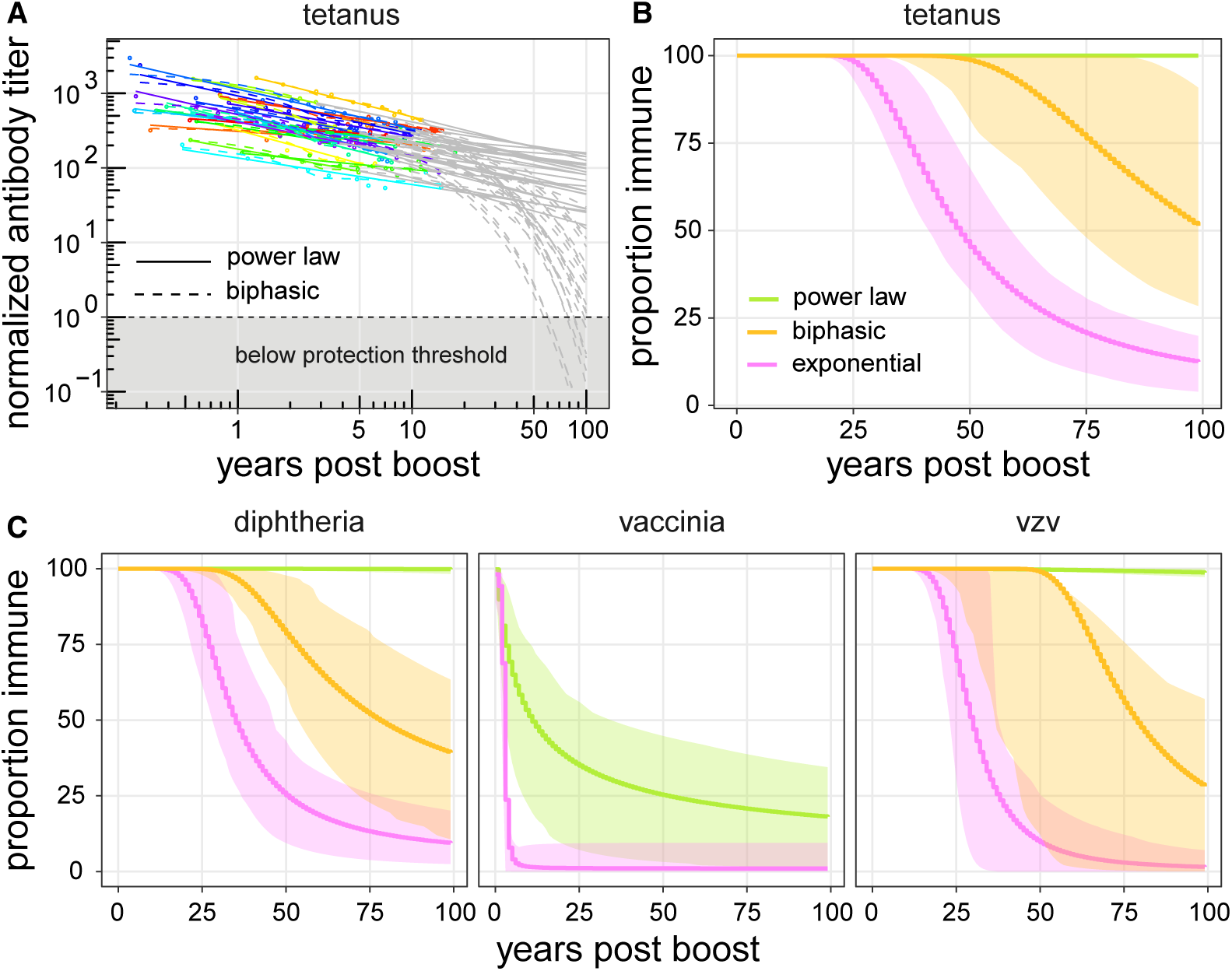
Model projections for time to loss of immunity. **(A)** The waning of immunity of individuals to tetanus as predicted by different models. Each individual is shown by a different color for the duration for the duration that the data was collected and extrapolations are shown in gray. The solid lines corresponds to a power-law model and the dashed lines to the bi-phasic model. **(B)** Predictions for the loss of protection at the population level over time for tetanus. The green line corresponds to the power-law model, the orange yellow line corresponds to the biphasic model and the orchid line corresponds to the simple exponential model. The colored shaded regions are 95% confidence intervals. We see that the power-law model predicts a much longer duration for antibody titer remaining above the threshold of protection than the exponential decay models. **(C)** Predictions of the models for the loss of protection at the population level over time for diphtheria, vaccinia, and VZV.

As shown in Fig 5, for the virus and vaccine antigens considered here, variation in the magnitude of the response (*c*_2_) has a larger effect on the longevity of protective immunity than changes in the power-law constant (*m*). This is expected because the different virus and vaccines have broadly similar values of the parameter *m* but very different values for the parameter *c*_2_. We also note that for most of the infections and immunizations in this study we expect lifelong protective immunity.

**Figure 5:**
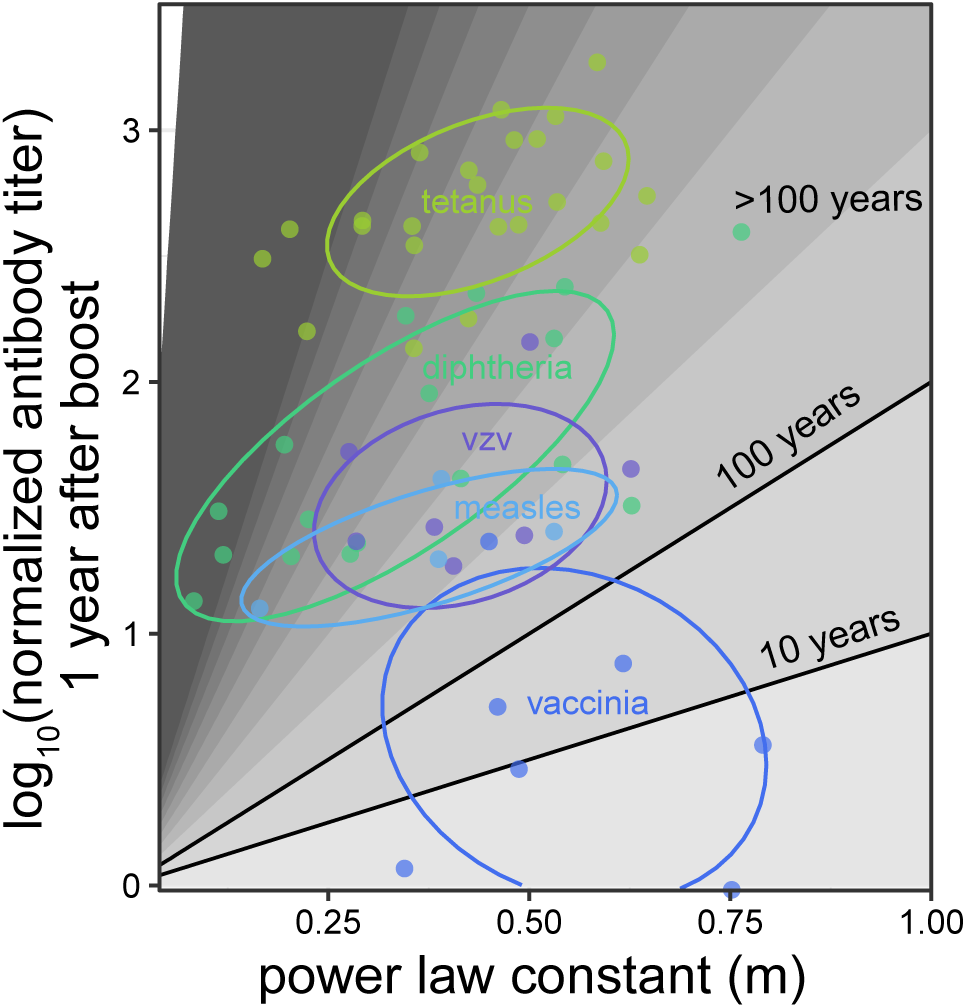
Summary plot. We plot how the magnitude of the response (*c*_2_ on the y-axis) and the power-law coefficient (*m* on the x-axis) determine how long antibody titers stay above the threshold for protection (represented with different scales of gray). Estimates for different individuals for different vaccines are points and the ellipses show 68% CI.

### The power-law allows estimation of rates of waning from short time-series

A major problem with estimating the longevity of immunological memory is that the long time-scale involved requires the collection of data many decades after immunization. This is particularly true for the exponential waning models where the rapid waning following the first 3 years after a boost was ignored and the rate of waning was considered only subsequent to this time [8, 13]. In contrast, the power-law model allows analysis of all the data on the waning of antibodies, including that from the first few years. In Table 2 we show that the power-law coefficients obtained from analysis of data from only the first 3 years following immunization or vaccination is very similar to that obtained if we use the entire timeseries, which for tetanus, diphtheria and VZV is about 10 years. We were not able to do this comparison for measles and vaccinia due to the paucity of long-term data (i.e., for periods greater than 3 years). In Fig 6 we compare the predictions for waning of immunity to tetanus when we use just the first 3 years of data vs the entire time-series. Predictions for other vaccines and viruses are shown in SI Fig S4. Consequently use of the power-law model might allow more rapid quantification of the waning of immunity compared with exponential waning models.

**Figure 6:**
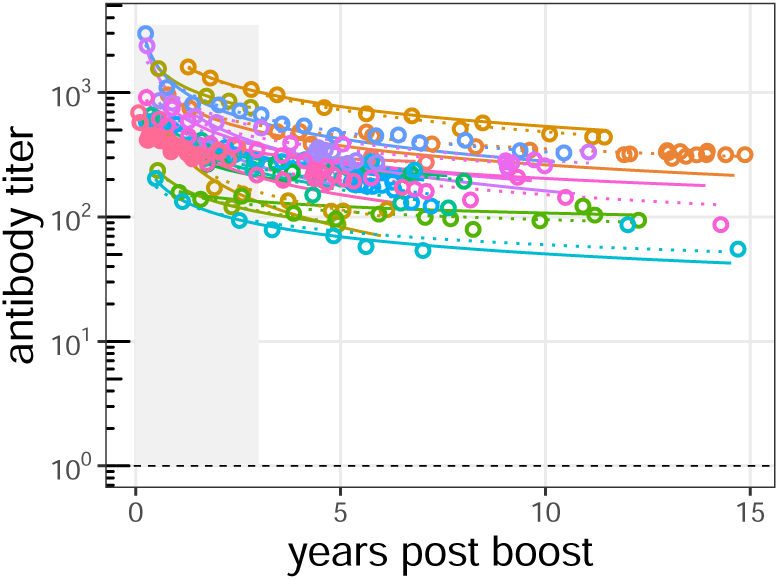
Prediction for waning to tetanus from data from year 0 to 3. We plot the prediction for waning of antibody for the entire timeseries using estimates obtained from data for first 3 years alone (solid lines) vs predictions using the entire timeseries (dotted lines). The black dashed line indicates the threshold for protective levels of antibody.

**Table 2:** Estimates of the power-law coefficient *m* using data from < 3 years post infection alongside estimates of *m* using the entire time series. Values in brackets are the SEM.

| Antigen | From $t < 3$ years | From entire timeseries |
| --- | --- | --- |
| Tetanus | 0.44 (0.06) | 0.43 (0.03) |
| Diphtheria | 0.39 (0.09) | 0.36 (0.05) |
| VZV | 0.42 (0.07) | 0.42 (0.05) |

## 3 Discussion

Our analysis of the waning of antibodies to a number of virus and vaccine antigens over a period of more than a decade allowed us to address whether the pattern of waning is better described by a power-law or exponential decay models. We found that a power-law model provides a better description and requires us to re-evaluate many current views and also suggests redesign of vaccine trials to measure the waning of antibodies.

Our findings suggest the time following immunization needs to be taken into account when comparing the rate of waning of antibodies to protein and toxoid vaccines, such as tetanus and diphtheria, with the waning of antibodies elicited by live virus infections such as measles, vaccinia and VZV. We suggest that earlier differences in estimates of rate of waning arose in large part because the waning of responses to tetanus and diphtheria were estimated relatively soon after vaccination when waning is relatively rapid. Our estimation of the duration of protective immunity to tetanus and diphtheria supports suggestions that additional adult boosters may not be required for toxoid vaccines [7, 10]. Our findings further suggest that previous estimates for the time it takes for antibodies to fall below a protective threshold needs to be revised upwards. The extent of the discrepancy for the estimates of the time to protection between the power-law and exponential models is greater when the magnitude of the response (compared with the protective threshold) is high, and when the estimates using the exponential model are from data at relatively early times after vaccination.

Our analyses have further found that the power-law model not only provides a better fit to antibody titer data, but also that a power-law model may potentially have a more practical advantage: it allows estimation of the long term decay of antibody titers from data for the waning of antibodies over the first few years after infection or immunization. Estimation of the parameters for power-law waning would benefit from measuring antibody titers at geometrically increasing time intervals after vaccination or infection rather than at equal time intervals. A disadvantage of using a power-law model, however, is that we need to know the time of infection or vaccination.

The power-law model we use is a simple model that describes the overall pattern of antibody waning with just 2 free parameters. Other, potentially more complex models have been suggested, including one which incorporates the generation of responses [23], and a modified power-law where antibodies fall only to a long-term level [14, 20, 26, 36]. One could also imagine other models that give rise to long-tailed functions for antibody waning. Here, we focused on a simple power-law model for a number of reasons including lack of data for the boosting phase of the response and varying amounts of data for the different virus and vaccine antigens (see Fig S5 in Supplementary Text). Further refinements to the power-law model will be facilitated both by more data of waning in individuals as well as by more nuanced understanding of the immunological basis of memory. Additional studies should include measurement of antibodies of different isotypes [37] as well as the patterns of waning for T cells [9, 38, 39]. A more nuanced and quantitative understanding of the cellular basis of immunological memory could also help formulate the term used for antibody decay. Exponential waning would be consistent with stochastic loss of plasma cells at a fixed rate in a time-independent manner akin to radioactive decay. The fact that this seems to not be the case can be attributed to heterogeneity in the rate of loss of plasma cells which is captured in the power-law model. Many processes might underlie this heterogeneity including different subpopulations of plasma cells [23, 40, 41], competition for preferred sites in the bone marrow [42], and progressive differentiation into longer lived cells [39, 43, 44, 45, 46]. The nature of the waning could also be affected depending on whether plasma cells are generated in extra-follicular or germinal center responses [47, 48, 49, 50]. Further studies should also take into account different levels of variation: within an individual host, between individuals, and among different pathogens.

### Limitations

We discuss the limitations of our current study of the waning of immunity, and further studies needed to overcome these limitations.

We consider the waning of IgG antibody titers in blood. Further studies are needed to understand the waning of immunity of different antibody isotypes and their titers in different locations. This would be particularly important for respiratory infections as these are controlled by IgA and to a lesser extent IgG1 isotypes that can migrate into the mucosal tissue where they are lost more rapidly following clearance of the infection [51, 52]. Understanding the dynamics of responses to respiratory infections such as influenza and coronaviruses is further complicated as they show rapid antigenic changes over time [53, 54, 55, 56].

Our analysis indicates that the time following immunization needs to be taken into account when comparing the rates of waning of antibodies to different virus and vaccine antigens, and on doing this we find that differences in the rate of waning of antibodies to protein toxoid vaccines and live virus infections is smaller than previously estimated. We do not imply that antibodies to all virus and vaccine antigens wane at the same rate. Indeed a few studies that compare waning to different virus and vaccine antigens at the same time after vaccination or infection suggest that there are differences in rates of waning [37, 57, 58]. Further studies will be needed to determine how the waning of antibodies depends on factors such as the degree of multivalency of the antigen and whether the response is primary or secondary [58, 59, 60, 61].

We use protective titers defined in the literature, and future studies could take a more nuanced approach by considering the different measures of immune protection, including changes in susceptibility to infection or pathology following infection [62, 63]. Additionally the current data is restricted to healthy adults and subsequent studies could consider how both waning and protection change with age.

## Conclusion

Despite major advances in our understanding of immunological memory many questions remain. We hope this work motivates future studies that give us a more nuanced understanding of immunological memory and how it depends on factors such as the type of infection (e.g. respiratory vs systemic [52]), the type of antigen and adjuvant [64], antibody isotype [41], following primary vs secondary exposure to new pathogens [61], and biological aging [65]. An integrated approach combining immunological and epidemiological observations with mathematical analysis may be needed to answer these questions.

## Data Availability

We are using public available datasets
All data produced in the present work are contained in the manuscript

http://www.plosbiology.org/article/info:doi/10.1371/journal.pbio.2006601#pbio.2006601.s006

## Acknowledgements

We thank Prof. Mark Slifka for his generosity both in sharing data and for discussions over many years. This study was funded by NIH grants U01 AI150747 and U01 AI144616 (RA, HA, AS and CH); and NIAID Centers of Excellence for Influenza Research and Response (CEIRR) contract 75N93021C00017 (KK).

## Supplementary materials

### Data used for analysis

**Figure S1:**
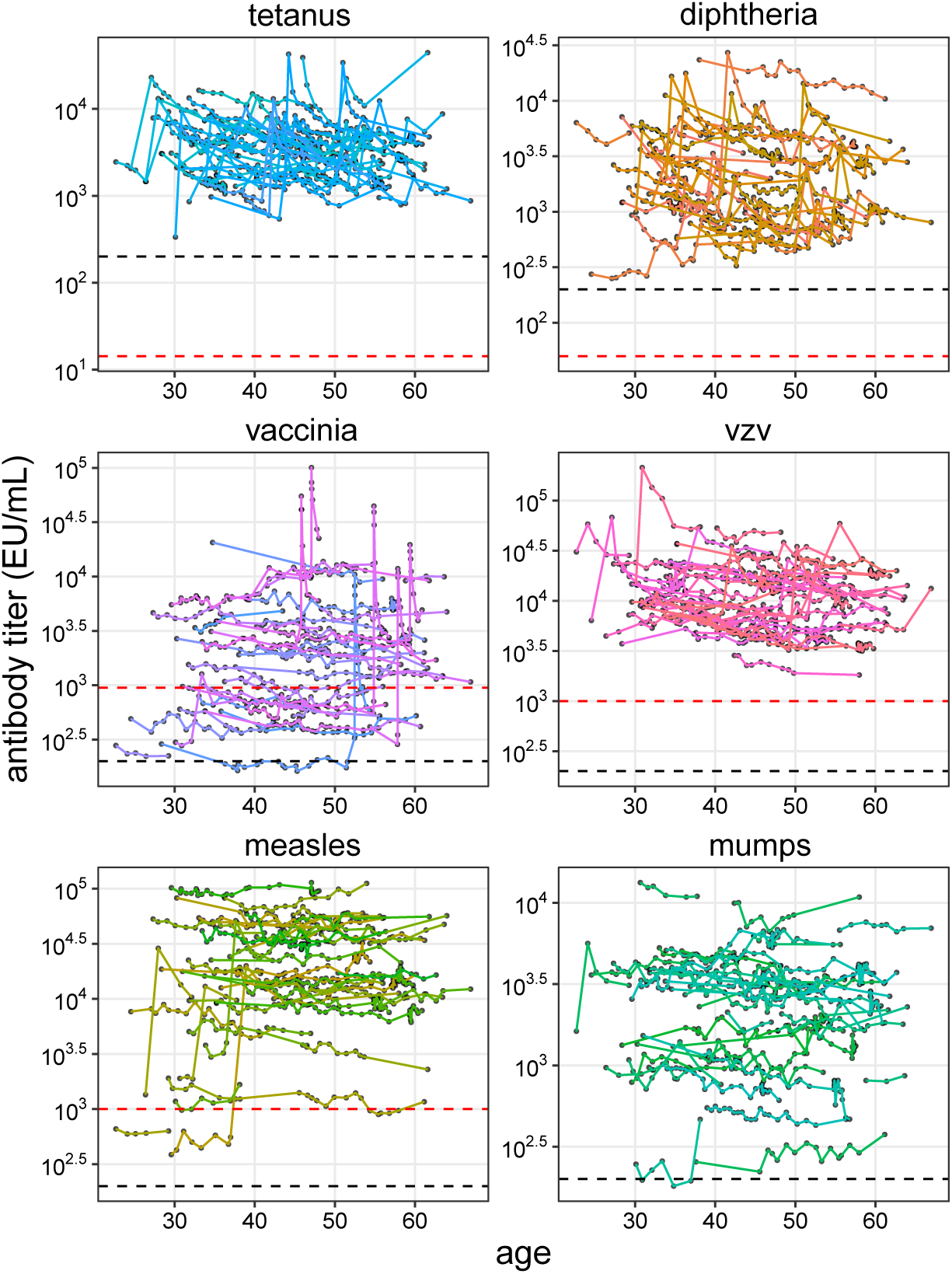
Raw dataset. Antibody titer (EU/ml) as a function of age for different vaccines are shown in different panels. In each panel, different lines joining the points represent different individuals’ time-series. The black dashed line in each panel is the sero-response level, and the red dashed line is the threshold for protection. For all vaccines, some individuals show spikes in their antibody titer followed by waning. Relatively few spikes are identified for live (attenuated) virus/vaccines (measles, mumps, vaccinia, and VZV) compared to toxoid based vaccines (tetanus and diphtheria).

**Figure S2:**
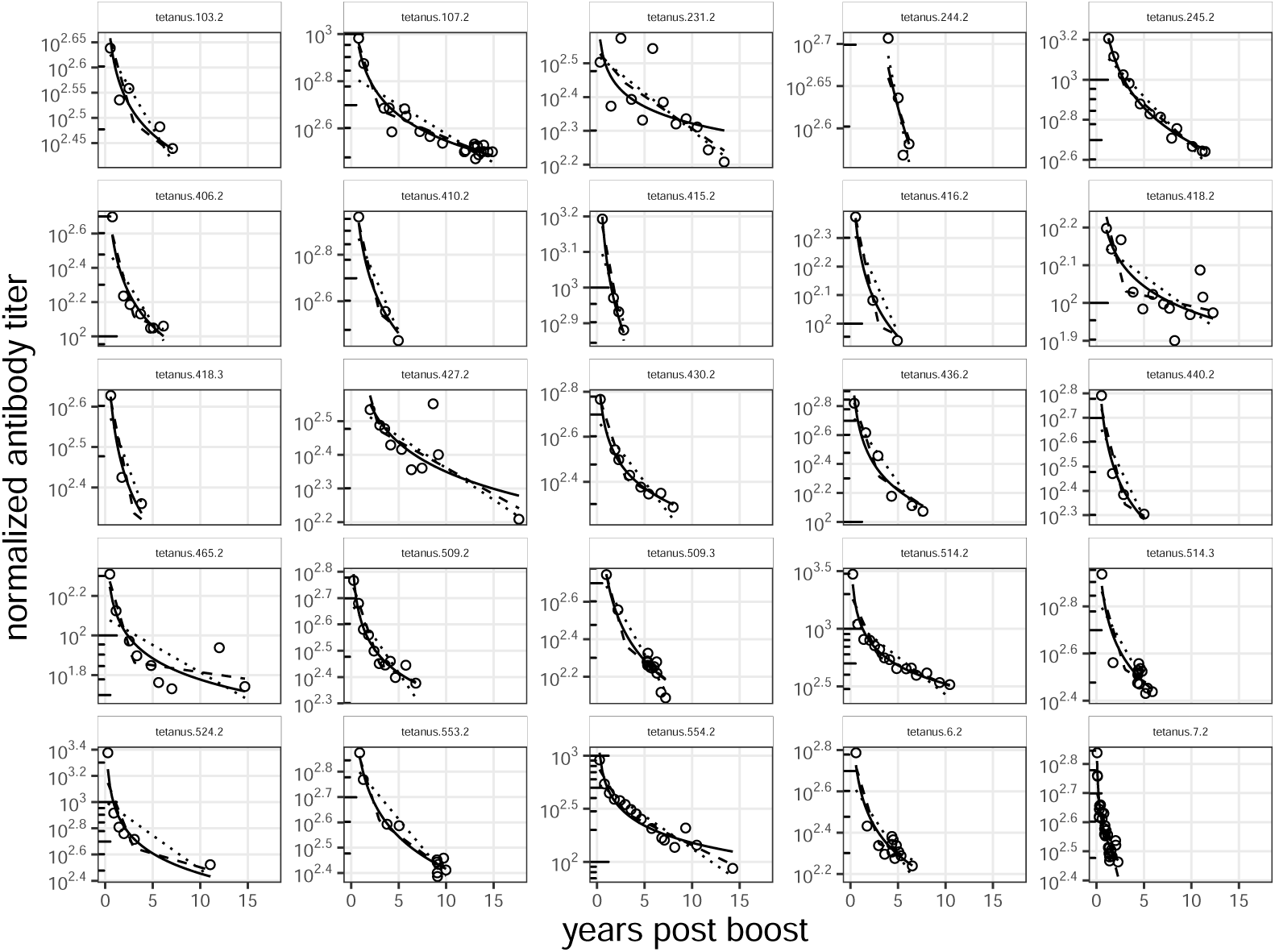
Exponential, bi-phasic exponential, and power-law model fits for the waning of antibodies to tetanus. Here we plot the antibody responses shown in Fig 1 in more detail with each individual in a separate panel.The fits for the exponential (dotted lines), biphasic exponential (dashed lines), and power-law model (solid lines) for tetanus are shown in different panels with log antibody titer axis and linear time axis. In each panel, points represent the data.

**Figure S3:**
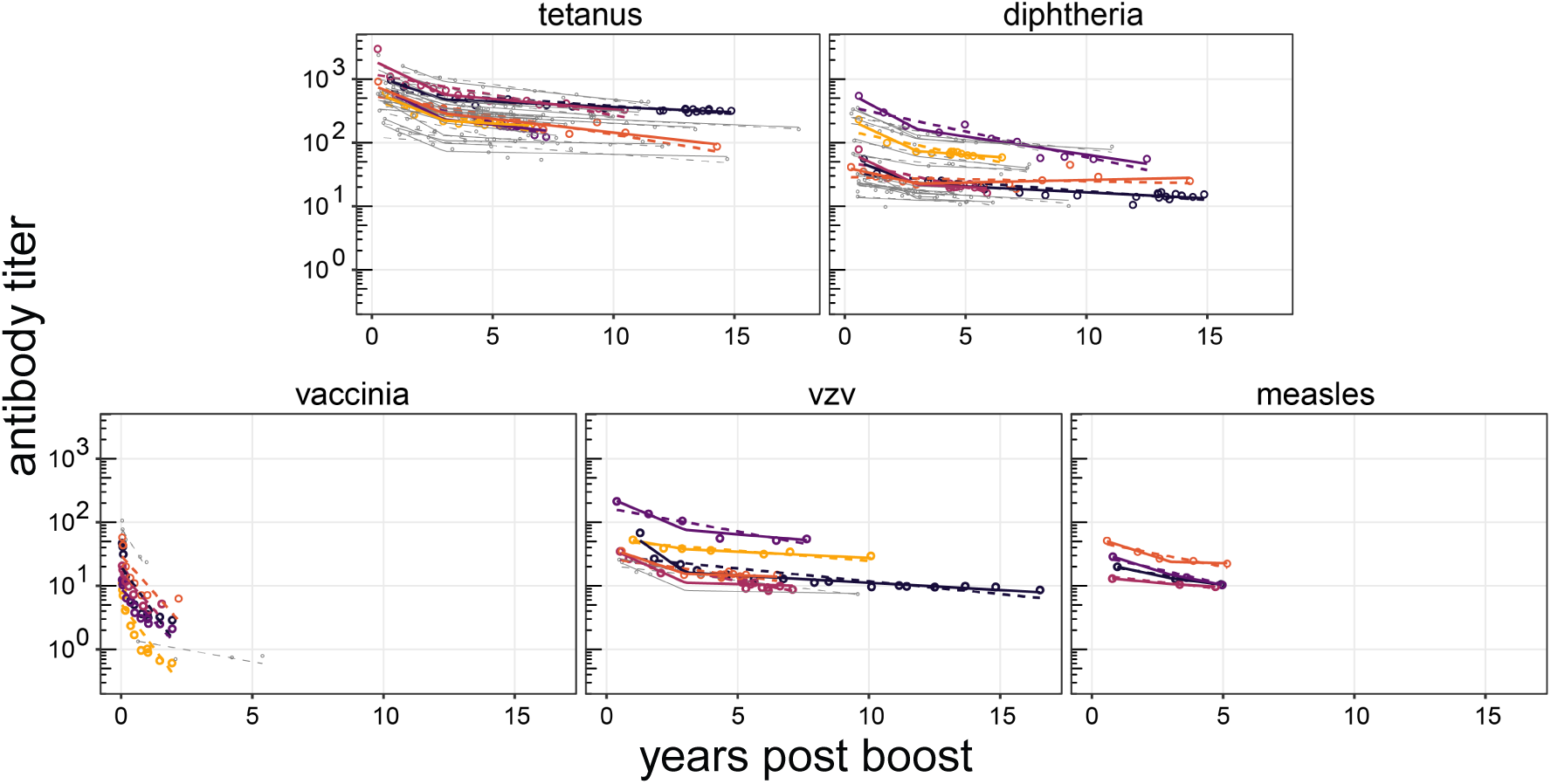
Exponential and bi-phasic exponential fits for all individuals for different vaccines. Individual fits for the exponential and biphasic exponential models for different vaccines are shown in different panels with log antibody titer axis and linear time axis. In each panel, points represent the data, solid lines and dashed lines represent the biphasic exponential and simple exponential model fits, respectively. 5 timeseries are highlighted in different colors for each vaccine. Others are colored as grey in the background. For vaccinia, the biphasic exponential model was not fit since all the individual time series were shorter than 3 years.

### Table S1. The half-life increases over time in a manner consistent with the power-law

The power-law predicts that the half-life increases linearly with time and this could account for much of the difference between previously estimates that suggested that antibodies to protein toxoid vaccines (tetanus and diptheria) wane faster than those to live virus infections and vaccinations (Vaccinia, VZV, and measles). This is shown in more detail in Table S1 below. We note that the longer half life calculated from data from adults for measles is higher than that predicted from the power-law model. Inspection of the data for measles reveals that about half the individuals appear to have slightly increasing antibody titers, despite no signs of a spike indicative of re-exposure, and if we restrict analysis to individuals with decreasing antibody titers to measles in adulthood then we get a similar half-life to the other virus and vaccine antigens considered in the table below. We do not currently know why some individuals showed increasing antibody titers to measles many decades after infection.

**Figure S4:**
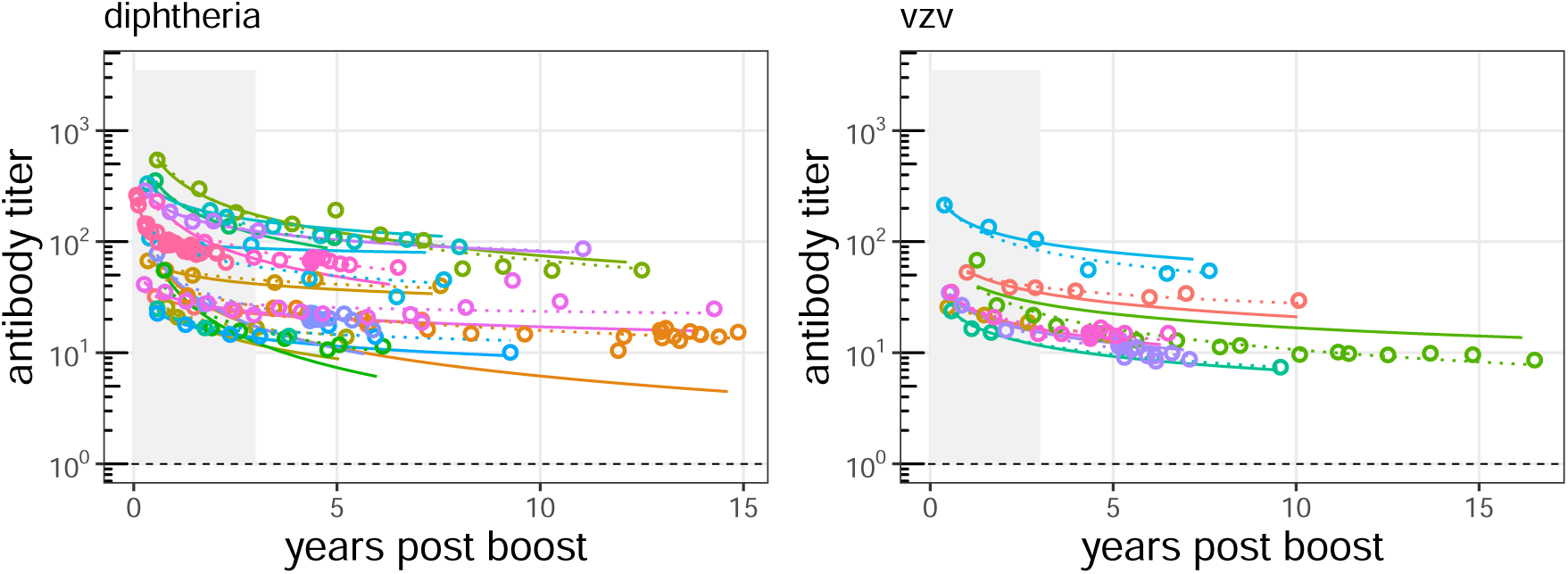
Prediction for waning for diphtheria and VZV. We plot the prediction for waning of antibody for the entire timeseries using data from the first 3 years alone (solid lines) vs predictions using the entire timeseries (dotted lines). The black dashed line indicates the threshold for protective levels of antibody.

**Table S1:** Half-life of decay for different vaccines at different time points. The instantaneous half-lives calculated as *T*_1/2_ = *Log*(2)/*k*. For the power-law *k* = *m*/*t*. Values and 95% CI shown. We see that the half-lives estimated by the power-law at about 8 years post vaccination are comparable to the estimates by Amanna et al. for tetanus and diphtheria and extrapolations for the half-life at 45 years are comparable to the estimates from Amanna et al calculated from data around this time post infection.

| Antigen | power-law model |  | Ammana (exponential) model |  |
| --- | --- | --- | --- | --- |
|  | Estimated at<br>~ 8 years | Extrapolated to ~<br>45 years | Estimated at ~<br>8 years | Estimated at ~<br>45 years |
| Tetanus | 13 (11-15) | 73 (64-86) | 11 (10-14) |  |
| Diphtheria | 15.5 (12-21) | 87 (68-120) | 19 (14-33) |  |
| Vaccinia | 10 (8-13) | 56 (46-72) | | 92 (46-∞) |
| VZV | 13 (10-17) | 73 (59 - 97) |  | 50 (30-153) |
| Measles | 15 (11-24) | 85 (62-133) | | 3014 (104-∞) |

### Some considerations of our choice for time of immunization

We use a simple power-law model where *t* = 0 is the time of vaccination or infection. As we do not have a precise time for vaccination we choose *t* = 0 as the midpoint between the datapoint just before and after a spike in antibody titer. We now consider the robustness of results to this approximation for the time of infection in Fig S5.

**Figure S5:**
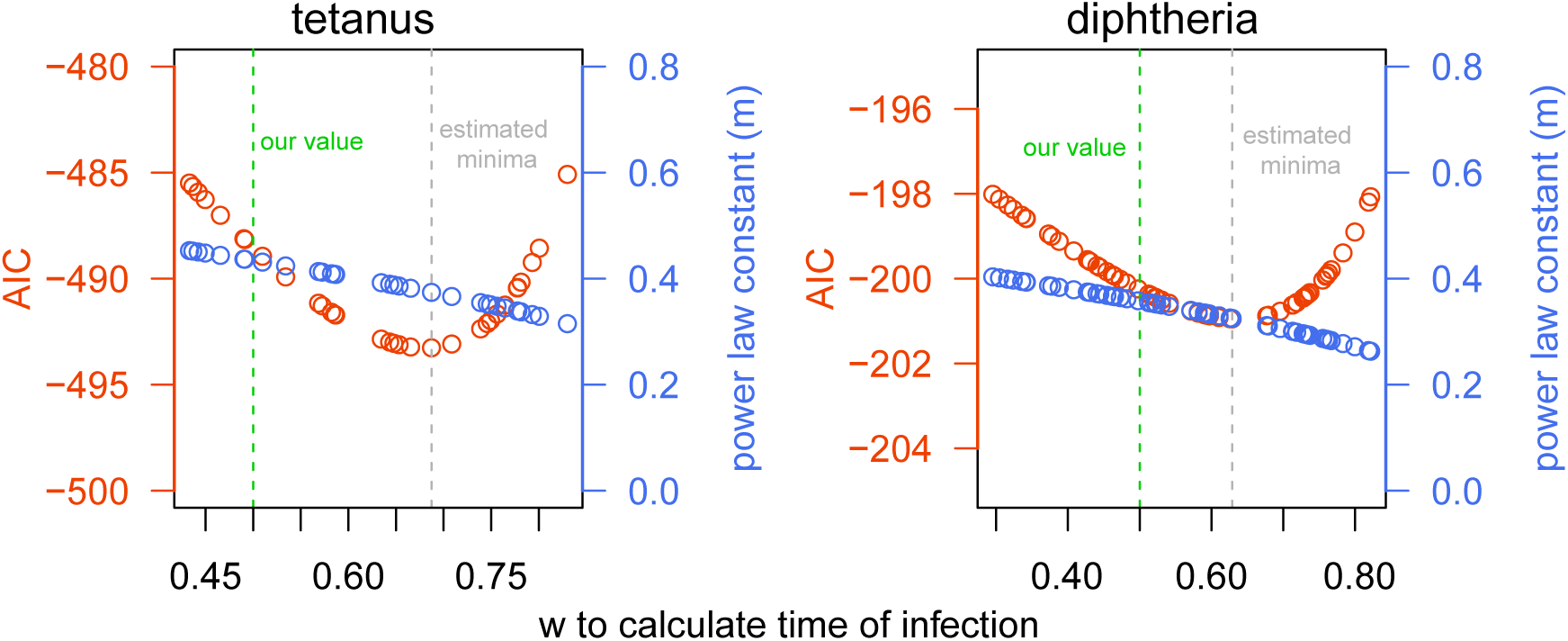
Effect of choice of the time of vaccination on the fits to tetanus and diphtheria. The time of infection or vaccination (*T* = (1 − *w*)*t*_1_ + *wt*_2_) was varied within the two time-points, *t*_1_ and *t*_2_ which the age just before and after the antibody spike. Thus *w* goes between 0 and 1 as we move the time of vaccination from the timepoints *t*_1_ and *t*_2_. The vertical green dashed line shows our choice of time for immunization (*w* = 0.5) and the grey dashed line the location of the minimum AIC value. We show that there is only a modest decrease in AIC and the value of *m* doesn’t change substantially between our value and the estimated minima, indicating the robustness of our assumption.

